# NUTRITIONAL IMMUNITY, ZINC SUFFICIENCY AND COVID-19 MORTALITY IN SOCIALLY SIMILAR EUROPEAN POPULATIONS

**DOI:** 10.1101/2020.11.04.20151290

**Authors:** Samer Singh, Amita Diwaker, Brijesh P. Singh, Rakesh K. Singh

## Abstract

The impact of Zinc (Zn) sufficiency/supplementation on COVID-19 associated mortality and incidence (SARS-CoV-2 infections) remains unknown. During an infection, the levels of free Zn are reduced as part of ‘nutritional immunity’ to limit the growth and replication of pathogen and the ensuing inflammatory damage. Considering its key role in immune competency and frequently recorded deficiency in large sections of different populations, Zn has been prescribed for both prophylactic and therapeutic purposes in COVID-19 without any corroborating evidence for its protective role. Multiple trials are underway evaluating the effect of Zn supplementation on COVID-19 outcome in patients getting standard of care treatment. However, the trial designs presumably lack the power to identify negative effects of Zn supplementation, especially in the vulnerable groups of elderly and patients with comorbidities (contributing 9 out of 10 deaths; up to >8000-fold higher mortality). In this study, we have analyzed COVID-19 mortality and incidence (case) data from 23 socially similar European populations with comparable confounders (population: 522.47 million; experiencing up to >150 fold difference in death rates) and at the matching stage of the pandemic (12 March - 26 June 2020; 1^st^ wave of COVID-19 incidence and mortality). Our results suggest a positive correlation between populations’ Zn-sufficiency status and COVID-19 mortality (*r*(23): 0.7893–0.6849, *p-*value<0.0003) as well as incidence [*r*(23):0.8084 to 0.5658; p-value<0.005]. The observed association is contrary to what would be expected if Zn sufficiency was protective in COVID-19. Thus, controlled trials or retrospective analyses of the adverse event patients’ data should be undertaken to correctly guide the practice of Zn supplementation in COVID-19.

## INTRODUCTION

Zinc (Zn) is a redox-neutral key micronutrient that plays an important role in immune competence and wellbeing (1-3). Worldwide, the explicit and implicit general recommendations for immune augmentation of the supposed ‘Zn-deficient’ population components and COVID-19 patients by healthy diet or Zn supplementation are in place. The ‘COVID-19 treatment panel guidelines’ of the Centre for Disease Control (CDC), USA, and others seemingly suggest Zn supplementation as an adjunctive therapy option ‘to complement the presumed protective role of Zn in infections and other immune and oxidative stress-related conditions/pathologies relevant to COVID-19 (4-7). A surge in the number of articles advocating a role of Zn supplementation in COVID-19 treatment/management has been observed. A search with keywords ‘((Zinc) AND (COVID-19)) AND (treatment)’ had yielded 117 articles in PubMed on 14 Jan 2021(8). Speculative benefit from Zn supplementation in COVID-19 has fueled its widespread prescription and over-the-counter purchases. This practice has gained momentum due to incorrect interpretation and implementation of CDC’s guidelines, inability to accurately assay Zn deficiency, and mild to no side effects of Zn overdose in healthy individuals. The actual figures of the total increase in Zn supplementation worldwide as a result of the COVID-19 pandemic are not in the public domain but from news articles, it may be surmised that it had already registered a surge by mid-April of 2020 and have steadily increased in different markets/countries since then (9-11). However, the clinical evidence supporting any beneficial effect of Zn supplementation or the ecological association studies supporting the potential positive impact of Zn sufficiency of the population/individuals on COVID-19 disease is nonexistent. Apparently, the push for Zn supplementation in COVID-19 is fueled by observed *in vitro* suppression of viral replication including that of coronavirus by high-doses of Zn [12, 13] which are physiologically irrelevant and would be unattainable *in vivo*. Globally, Zn deficiency among populations ranges from 3-54% (14). The developed European and North American nations generally have the highest sufficiency while African and Asian nations have the least. Its deficiency in populations is widely associated with vulnerability to infectious diseases, chronic immune system disorders, and other medical conditions (3-5,7,14,15).

The Zn levels are tightly regulated in the human body and its differential levels are maintained in various organs, tissues, and extracellular milieu including serum through a complex homeostasis mechanism involving multiple regulators and transporters (1, 16-18). Optimal levels are maintained in different tissues, organs as well as immune cells to regulate inflammation and pathogen clearance, reduce reactive oxygen radicals, and affect various bodily processes including lipid metabolism and glucose levels. During infections, the levels of Zn are reduced in serum and it is redistributed to tissues and immune cells as a part of ‘nutritional immunity’ to enhance immune cells’ function and check pathogens’ growth and replication (1, 16, 18, 19). The intracellular Zn reserves are also mobilized and reallocated in the presence of intracellular pathogens in an attempt to reduce the availability of free Zn to limit their survival and replication potential. Simultaneously, it signals the neighboring cells about the presence of pathogens and induces a balanced inflammatory response (REFs in 1, 2, 16, 18, 19). Zn insufficiency is associated with frequent infections and chronic inflammation. Due to the unavailability of accurate measures (assays) to diagnose Zn deficiency, serum levels along with relevant deficiency symptoms are used to prescribe Zn supplements in pathologically deficient individuals. Nonetheless, Zn supplementations are known to promote adverse effects in several disease conditions which are currently also identified as comorbid conditions for COVID-19 patients and frequently observed in the elderly, *e*.*g*., sickle cell disease, chronic kidney diseases (CKD), cardiovascular diseases (CVD), and coronary heart disease (CHD) (15,20-23).

We hypothesized if higher Zn levels would be playing the supposed protective role against COVID-19, the COVID-19 associated mortality (and incidence) would negatively covary with the Zn sufficiency of populations. The European populations that experienced the 1^st^ wave of SARS-CoV-2 infections simultaneously and had more similar confounders, such as, high Healthcare Access and Quality (HAQ) Index, life expectancy, median age (24,25), etc. as compared to other parts of the world, offer an excellent opportunity to test such assertions. These relatively similar populations had been also previously analyzed for proposing other potential protective variables (26-31) and variously referred to as socially similar. In the current study, first waves’ phase-matched pandemic data of socially similar European populations with supposedly comparable confounders were analyzed to test the null hypothesis that Zn sufficiency is not correlated with COVID-19 associated mortality (and incidence).

## MATERIAL AND METHODS

### Study populations and Zn sufficiency status

The COVID-19 incidences and mortality data of twenty-three socially similar European populations (Total 522.47 million) with similar confounders (26-30) were collected from Worldometer’s coronavirus webpage (24). The data for the 1^st^ wave of SARS-CoV-2 infections in these countries (12 March to 26 June 2020) are presented as COVID-19 cases per million (CpM) and Deaths per million (DpM) in Table 1 (see Supplementary Table 1 for extended dataset up to 26 August 2020). The populations’ Zn sufficiency/deficiency level estimates were from Wessells and Brown, 2012 (14). For an overview of the worldwide country-wise Zn sufficiency of the populations and COVID-19 impact refer to Supplementary Figure 1. The notification rates of COVID-19 deaths and incidences in different socially similar European populations during the passage of SARS-CoV-2’s 1^st^ wave-of-infections are provided in Supplementary Figure 2. The European countries with comparable confounders and at a similar phase/stage of the pandemic were selected for the current analysis to arrive at more dependable conclusions (26,28,29,31).

**Table 1.**
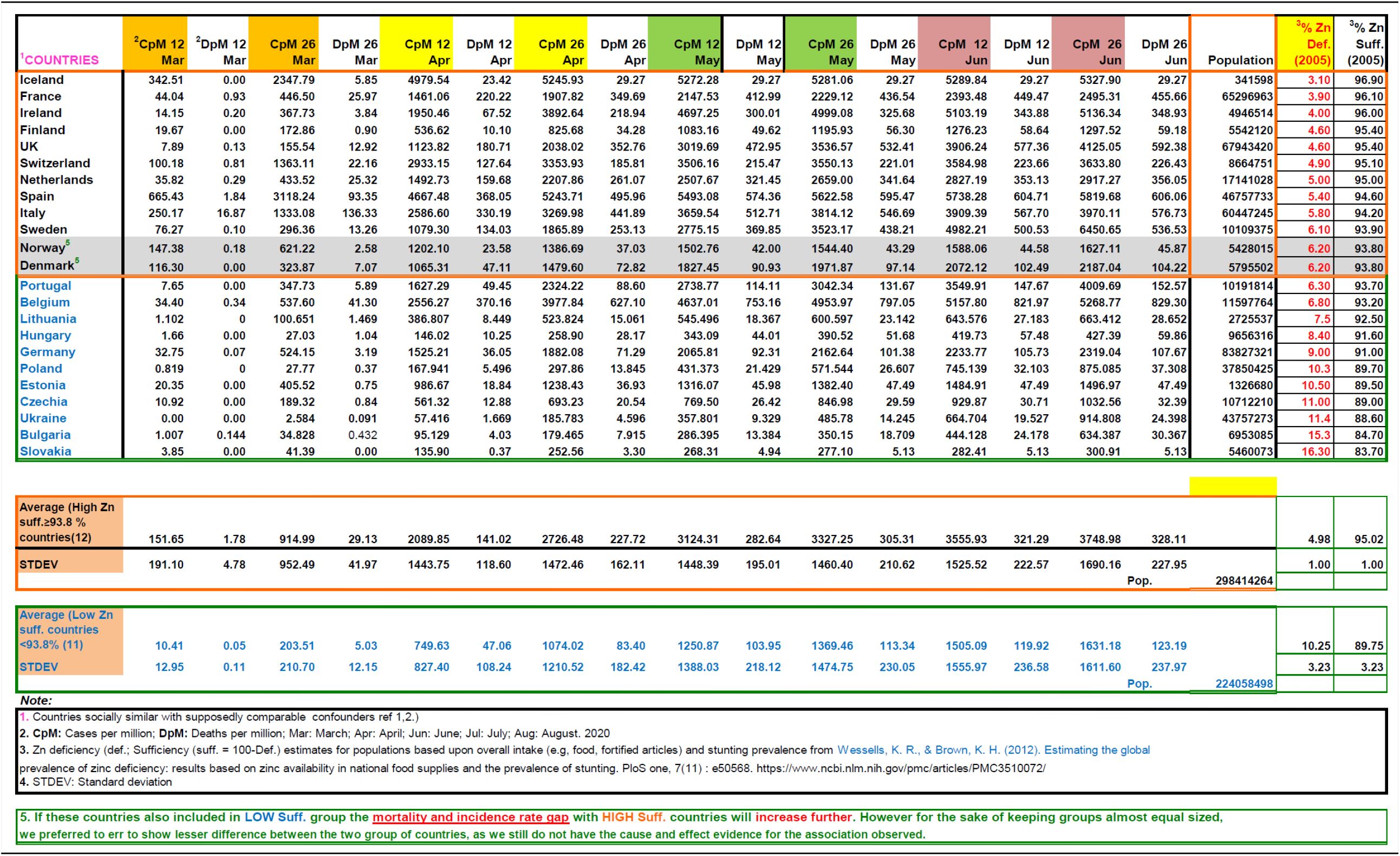
COVID-19 incidences and mortality in socially similar European populations.

### Statistical analysis

In order to examine the relationship between the prevalence of Zn sufficiency and deaths in the populations due to COVID-19, we obtained a Pearson correlation coefficient that provides the degree of relationship in real number, independent of the units in which the variables have been expressed and also indicates the directionality of the relationship, *i*.*e*., positive or negative. The formula for the Pearson’s correlation coefficient is as follows

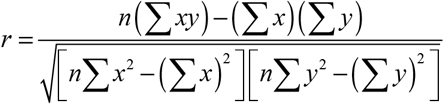; where *x* and *y* are the variables of interest and *n* is the number of paired observations.

We aimed to fit several statistical models including both linear and non-linear models for the empirical data. Various types of models were tested and among these, the best-fitted models have been considered here. For non-linear models, polynomials have been considered which can be mathematically expressed as:

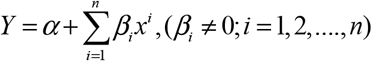

Where *α* is the constant, *β*_*i*_*’s (i=1,2,…*.*n)* are the regression coefficient of *X*_*i*_*’s (i=1,2,…*.*n)* and *X* is the independent variable, i.e., Zn sufficiency and *n* is a positive integer referred to as polynomial of degree *n*. For *n*=0, it becomes a constant function, whereas for *n*=1, it is a polynomial of degree 1 (i.e., a simple linear function), and for *n*=2 it is a polynomial of degree 2 (i.e., quadratic polynomial). For different degrees of polynomials the models employed can be simply expressed as under:

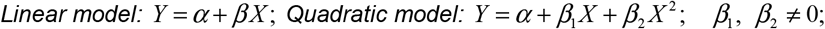

For the exponential curve, the relationship is mathematically expressed as *y* = *a e*^*β x*^. The explanation of various parameters (coefficients) of the models is that for a unit change in *‘X’* we observe a change in ‘*Y’* equal to the value of respective coefficients. All the computations were performed in Microsoft Excel. COVID-19 mortality and incidence, without applying extra exclusion/inclusion criterion of the risk factors or supposed confounders (e.g., age, sex, age distribution, comorbidities, populations’ density, etc.), were assessed for correlation with populations’ Zn sufficiency, as done previously (26, 29). Interim case fatality ratio (i-CFR) for the populations/groups were calculated as ‘(reported deaths/reported cases)*100. P-value <0.05 was considered statistically significant.

### Epidemiological map display and required color manipulation

Global maps showing country-wise Zn levels, as well as COVID-19 incidence and deaths, are adapted from (25). The Choropleth maps displaying the variation of variables for the selected countries were created using ‘Inkscape’ software.

## RESULTS

The socially similar countries selected for the analysis had been more severely affected by the COVID-19 as compared to other parts of the world despite being some of the least Zn deficiency countries (Supplementary Figure 1). Similarly, the nations of North America with higher Zn sufficiency levels were also found to be more severely affected by COVID-19 as compared to other parts of the world with lower Zn sufficiency. The European nations included in the current analysis were almost at a similar stage of pandemic (Supplementary Figure 2). The nations included in the current study had registered only 15.71% cases (1552 thousand cases) of the total global cases (incidences) by 26 June 2020 but about 34.2% of total global deaths (173 thousand deaths). As the wave of SARS-CoV-2 infections spread to other parts of the world by 26 August 2020, the relative contribution of the selected countries changed to 8.85% of cases and 21.78% of total global COVID-19 deaths. Nevertheless, the mortality rate remained disproportionately high in countries with better Zn sufficiency. The correlation (Pearson’s) and regression analysis were performed on these twenty-three socially similar European populations’ Zn sufficiency levels and COVID-19 incidences and mortality data (12 March to 26 June 2020) without considering any additional exclusion/inclusion criterion. The observations made are briefly summarized below:

1. ***Globally COVID-19 mortality rates positively correlate with Zinc sufficiency levels*:** Zn sufficiency/deficiency levels vary across the globe (Supp. Fig. 1). Generally, differential COVID-19 mortality rates had been observed among countries. The European and North American countries with higher HAQ index, life expectancy, median age (24, 25) came out as the worst affected. Previously, trained-immunity (26, 27), BCG vaccination (28), vitamin D (29), Zn deficiency (30) had been associated with reduced COVID-19 mortality using early-stage pandemic data (26-30). They had also attracted criticism for the inappropriateness of the data set, inclusion/exclusion criteria, non-matched pandemic stages among countries, and more importantly the loss of association by mid- or post-peak-of-infections (REFs in 25, 27, 29, 31). Globally the populations were exposed to COVID-19 at different time points, had different lifestyles, social structures, preventive measures in place, and access to medical facilities. So, the supposed associations drawn from them could be tenuous due to disparate data sets. To arrive at more dependable conclusions, when the pandemic phase-matched (Supp. Fig. 2) populations with comparable confounders (26,28,29,31) were analyzed for the COVID-19 mortality rate in socially similar populations [Boxed North America (NA) and European Union (EU)], it strongly associated with their Zn sufficiency (Supp. Fig 1; time-series graphics available at reference 25). Incidentally, the countries of Europe, North America, and South America that have reported high mortality have been also the high Zn sufficiency countries.
2. ***Zinc sufficiency strongly and significantly correlated with COVID-19 mortality in socially similar countries:*** The Zn sufficiency in twenty-three socially similar European populations (Fig 1A, Table 1) was positively associated with reported COVID-19 deaths and cases per million (Fig. 1B-C). The COVID-19 associated mortality rate (deaths/million) and incidence rate (cases/million) covaried exponentially with Zn sufficiency of the populations (Fig. 1D-E). The populations with Zn sufficiency >92.5% had experienced up to 9X higher deaths per million population (Fig. 1D). Pearson’s correlation for the synchronized period of reported COVID-19 cases and deaths among the countries (26 April to 26 June, Supp. Fig. 2), was strong [Table 2, Deaths per million: *r*(23)= 0.7455–0.6849, p<0.0003; Cases per million: *r*(23)= 0.8084–0.7629, p<0.00002]. The division of countries using >93.7% sufficiency cutoff, indicated high mortality (5.79 to 2.66X) and incidences (4.5 to 2.3-fold) in high Zn sufficiency populations (n=12) as compared to low sufficiency populations (n=11). Some of the outlier populations that fared extremely well and displayed quite low mortality among the higher Zn sufficiency countries, *e*.*g*., Iceland, Finland, Norway, Denmark, also happen to have higher levels of ‘trained immunity’ and serum ‘vitamin D’ levels - two other protective variables proposed earlier (26, 27, 29).

**Fig. 1.**
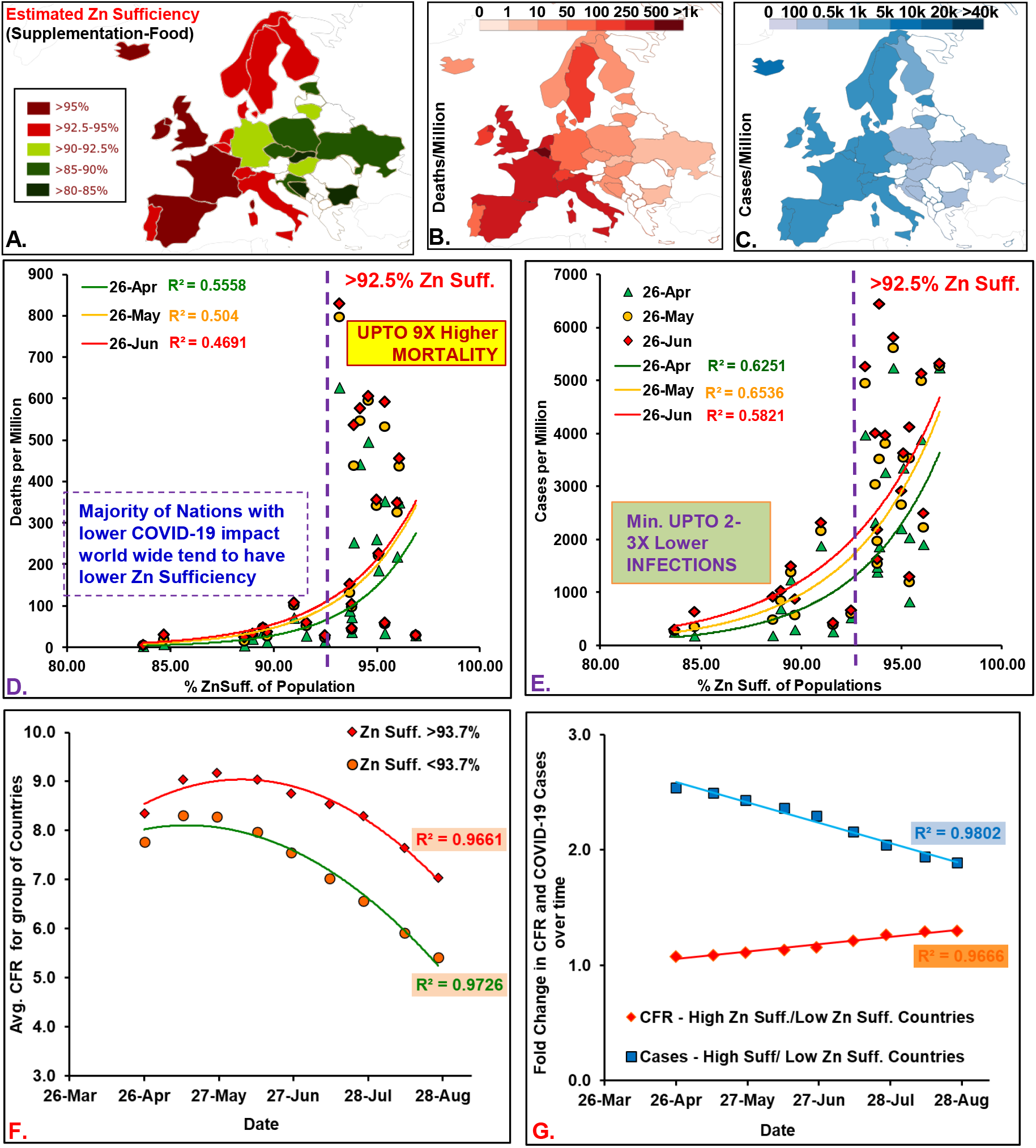
Zinc sufficiency of the socially similar European populations correlated with COVID-19 mortality. Study populations: **(A)** Zn sufficiency estimates, **(B)** and **(C)** deaths and cases/million (26^th^ April). [Note: Global maps [18] displayed color scheme variations performed using ‘Inkscape’]. Deaths **(D)** and cases **(E)** per million population positively correlated with their Zn sufficiency levels. **(F)** For the group of countries with different Zn sufficiency (suff.) the interim case fatality ratio [i-CFR or Avg. CFR = (reported deaths/ reported cases) *100] remained different throughout the period after peaking in May 2020. **(G)** Fold change in the relative CFR for high suff. with respect to low suff. countries have been registering a steady increase while the gap for cases has kept shrinking.

**Table 2.** **Correlation Analysis of COVID-19 mortality and incidence with Zinc sufficiency of populations**. Correlation analysis for the most synchronized period of SARS-CoV-2 infections in twenty-three socially similar European countries (highlighted yellow period – covers the rise and flattening of COVID-19 case and mortality rate (see Supp. Fig. 2). P-value <0.05 is referred to as statistically significant.
3. ***Higher Zn sufficient socially similar countries/populations consistently reported higher COVID-19 case fatality rates (CFRs) and incidence rates:*** The CFRs for the group of countries with higher Zn sufficiency (>93.7%, 12 countries) remained higher than that for the group of countries with lower Zn sufficiency (<93.7% 11 countries) throughout the study period (till 26 August 2020; Fig. 1 F). It started to decline post-peak-of-infections, *i*.*e*, 12 May 2020. This could be reflective of more uniform reporting in later stages of the pandemic or decreased load on medical infrastructure resulting in better patient care and lower adverse outcomes. The relative difference in the COVID-19 cases per million between two groups of countries decreased over time but the rate of CFR decrease had been slower in high Zn sufficiency populations than that in lower Zn sufficiency populations, resulting in a progressively increasing CFR gap between the two sets of countries during the study period (a CFR gap of 16% on 26 June become 30 % by 26 August 2020) (Fig. 1G).

| **CORRELATION ANALYSIS** |  |  |  |  |  |  |  |  |
| --- | --- | --- | --- | --- | --- | --- | --- | --- |
| Data-Date | 12 Mar | 26 Mar | 12 Apr | 26 Apr | 12 May | 26 May | 12 Jun | 26 Jun |
| Average Deaths/Million Pop. ± STDEV | 0.95 ± 3.50 | 17.61 ± 33.16 | 96.08 ± 121.08 | 158.70 ± 183.58 | 197.18 ± 221.29 | 213.49 ± 236.32 | 224.98 ± 246.55 | 230.11 ± 250.35 |
| DEATHS/Million(Log) vs Zn SUFF. - Correlation $r(23)$: | 0.3731 | 0.7893 | 0.7642 | 0.7455 | 0.7213 | 0.7099 | 0.6989 | 0.6849 |
| $p$-value | 0.0795 | 7.6E-06 | 2.19E-05 | 4.5E-05 | 0.0001 | 0.0001 | 0.0002 | 0.0003 |
| Average Cases/million Pop. ± STDEV | 84.10 ± 153.42 | 574.71 ± 778.36 | 1448.88 ± 1349.80 | 1936.17 ± 1569.23 | 2228.32 ± 1685.30 | 2390.92 ± 1747.82 | 2575.09 ± 1833.38 | 2736.12 ± 1943.94 |
| CASES/Million (Log) vs Zn SUFF. - Correlation $r(23)$: | 0.5658 | 0.6513 | 0.7681 | 0.7906 | 0.8067 | 0.8084 | 0.7966 | 0.7629 |
| $p$-value | 0.0049 | 0.0008 | 1.88E-05 | 7.1E-06 | 3.33E-06 | 3.05E-06 | 5.42E-06 | 2.3E-05 |

## DISCUSSION AND CONCLUSION

Overall a general positive correlation of COVID-19 mortality and cases of populations with their Zn sufficiency levels was observed during the 1^st^ wave of SARS-CoV-2 infections, which sustained even on flattening of the curves. The socially similar European populations with comparable potential confounders which were also at a similar stage of the pandemic and had reported more than 399 cases per million population were included in the correlation analysis. This group of high COVID-19 impacted countries globally accounted for 34.2% mortality (173 thousand deaths) by 26 June 2020, giving confidence to the observations made. The correlation of Zn sufficiency of the study populations with COVID-19 mortality and cases for the whole study duration was similar to that was observed previously with an earlier phase COVID-19 pandemic data of a smaller set of countries [30]. The correlation remained stable and statistically significant for the whole study period encompassing the relevant phase of the wave-of-infections, unlike other risk factors or protective variables proposed previously using early phase pandemic data (27,29,31). The association observed in the current study is even stronger than those being actively discussed, debated and evaluated in clinical trials (28, 29, 31). It may be suggested that the association observed warrants a thorough investigation to arrive at a more meaningful conclusion about the role of Zn supplementation in COVID-19 outcomes.

It would be pertinent here to discuss the limitations of the data set used for the current analysis. The COVID-19 data had been collated by different agencies and had been subject to adjustments from time to time. Depending upon the aggregators, there also exist some minor differences between different COVID-19 databases, e.g., during 1^st^ wave of infections the maximum cases reported per day by Ireland as per ‘https://coronavirus.jhu.edu/map.html’ were 1515 cases on 10/04/20, while it was 1508 as per ‘Worldometer’. Similar size adjustments of the data could affect the early-stage data analysis more drastically than the adjustment made to later-stage data - making the early-stage associations inherently less reliable. The Zn deficiency/sufficiency estimates for the populations used in the current analysis are for the year 2005 that was published in 2012 (14). The estimate was based upon the food balance sheets, the bioavailability of Zn in the food supply, estimated physiological requirement of Zn in individuals, population demographics, and stunting observed in children. The revision of the Zn deficiency/sufficiency estimates for populations is long overdue. Since the estimates published by Wessells and Brown, 2012 remain the latest, they had been used as an approximation for the current levels of populations. However, for the sake of current analysis and discussion, it may be assumed that drastic negative changes in Zn sufficiency levels have not happened among these countries and the overall relative Zn sufficiency levels would have been maintained. The general improvement in the development index and the availability of diverse foods and supplements along with reduced stunting in children in the last two decades as per data available with the Food and Agriculture Organization (FAO) of the United Nations (UN) and World Health Organization (WHO) further strengthens the notion. The analysis presented in the current study may need to be revisited whenever more accurate Zn sufficiency/deficiency estimates of the populations for the current period become available. The availability of COVID-19 patients’ data concerning Zn supplementation and dietary food intake during COVID-19 is expected to provide the most accurate picture of the association.

The Zn bioavailability reduction during infections is a common component of ‘nutritional immunity’ (1, 16, 19, 32). The drop in serum Zn levels is primarily attributed to a proinflammatory cytokine-mediated alteration in the expression of hepatic Zn transporter proteins during the acute phase of inflammation through squelching of serum Zn coming from the diet (33). The cell-mediated Zn restrictions, reallocation, and mobilization (primarily mediated by ZIP family of transporters, *e*.*g*., ZIP8 in *Mycobacterium tuberculosis* (34), as well as extracellular Zn sequestration (primarily mediated by S100 family of proteins, e.g., calprotectin, psoriasin, calgranulin, etc.), is supposed to be part of an elaborate intricate immune mechanism to both starve and/or intoxicate pathogen with this key element as per differential need (REFs in 1, 2, 18, 19). Calprotectin, an S100A8/S100A9 heterodimer, seems to be a key player in immune regulation through its ability to chelate Zn and other metal ions (1, 35) and also serve as a damage-associated molecular pattern (DAMPs) and a ligand for Toll-like receptor 4 (TLR4) (36). It can account for >40% of cytoplasmic proteins in neutrophils during infection (37). *In vitro* studies have shown an important role of calprotectin in inhibiting the growth of the pathogens while the supplementation of Zn has been shown to revert the calprotectin-mediated growth inhibition (38-40). Here the Zn starvation strategy could primarily interfere with the Zn-dependent infection process and replication while the intoxication could help the generation of ROS in a localized manner for effective pathogen incapacitation and elimination (REFs in 1, 2, 32). The existing pieces of evidence point to the mobilization of Zn during infection as a measure to not only limit its availability to invading pathogens but also prime immune cells to kill pathogens.

Systematic reviews of Zn supplementation studies had shown a significant reduction of diarrhea incidents and the associated deaths in Zn deficient children (REFs in 41-43). It now forms a part of global diarrhea control measures. Several studies using a small set of individuals/patients do indicate a benefit of Zn supplementation in other diseases as well including pneumonia and lower respiratory tract infections which are relevant to COVID-19 (REFs in 41). However, the systematic review and meta-analysis of the previous studies do not support the supposed positive influence of Zn supplementation on the prevention and treatment of different diseases when Zn sufficient individuals were also included in studies (REFs in 41,43). Though Zn sufficiency promotes the well-being and integrity of respiratory cells during lung inflammation and protects them from injury arising from respiratory tract infections (RTI) and decrease the incidence of infections by regulating immune functions and response (44), the ability of Zn supplementation to help reduce the infection, severity or treatment of lower RTI (a comorbid condition in > 50% COVID-19 deaths) has not been unequivocally supported in different Zn sufficiency patient populations. A Cochrane Systematic review that summarized 73 studies of Zn supplementation performed in Zn-sufficient and deficient children (low and middle-income countries) up to the age of 12 years did not find evidence for the reduction in RTI (prevention) but indicated a negative effect on Cu status (42). However, when the same set of studies additionally considered the iron supplementation status a reduction in death risk ratio from RTI was observed as per the studies summarized in the WHO document available in e-Library of Evidence for Nutrition Actions (eLENA) [43]. Similarly, as per studies summarized in WHO guidelines for tuberculosis patients, Zn supplementation does not have desired beneficial effects on tuberculosis patients who chronically have lower circulating Zn levels (45). A more recently updated meta-analysis of Zn micronutrient supplementation also could not find augmentation of the tuberculosis therapy outcomes as assessed at different time points (*e*.*g*., sputum-culture or sputum-smear positivity, chest x-ray clearance; death in HIV positive patients) (46). The findings of a ZINC trial, recently reported in JAMA, that evaluated the effect of Zn supplementation on survival of HIV patients – who have also suffered during the COVID-19 pandemic, were discerning (47). A detrimental effect of Zn supplementation on the overall survival of study participants was observed. The study evaluated the effect of Zn supplementation on markers of mortality, the risk of cardiovascular disease, the levels of inflammation and associated microbial translocation, and HIV disease progression among HIV-positive alcoholic veterans. It concluded that Zn supplementation did not improve any of the assessed markers among 69% of the participants (i.e., survivors) who completed the 18-month study period. A closer look at the cause of dropouts as provided in the supplementary table (eTable. ‘Causes of Death in the ZINC Trial by Randomized Group’) surprisingly provides a different narrative. It highlights a potentially serious detrimental impact of Zn supplementation on a section of the HIV-infected cohort. The Zn supplemented group in comparison to the control group had observed almost two-fold higher all-cause mortality, *i*.*e*., 21/126 *vs* 12/128. If we remove cases with the unknown cause of deaths from both groups, the deaths were about 3 times more frequent in the Zn supplemented group (19/126 vs 7/128). Distribution of deaths in the Zn supplemented *vs* control group concerning the COVID-19 relevant conditions (23) was more surprising, *e*.*g*., HIV-related deaths: 7/21 *vs* 2/12; overdose-related deaths 4/21 *vs* 1/12; deaths from pneumonia: 3/21 *vs* 0/12; deaths from suicide: 2/21 *vs* 0/12; deaths from Thromboembolism: 1/21 *vs* 0/12.

Two notions that are supposedly contributing to the increase in Zn supplementation for COVID-19 management are, a) Zn can block viral replication, b) patients reporting COVID-19 symptoms have lower serum Zn levels. It must be remembered here that the free Zn levels required to achieve viral replication inhibition *in vivo* are not possible without killing the host (12). During infections and fever as a result of the natural ‘nutritional immunity’ response, the availability of cellular as well as the systemic Zn (from circulation) is reduced to impede the growth of pathogens (1, 2, 19, 48). The redistribution of Zn from circulation is part of a mechanism to limit the oxidative damage to peripheral tissues, away from circulatory or other vital systems during infections, which may experience a transient or sustained localized increase of oxidative radicals as a part of an immune response.

The serum Zn just represents <0.2 % of total Zn present in the body. There is a relative dearth of studies characterizing the effect of Zn supplementation on the kinetics of Zn serum levels during acute viral infection when the acute phase response actively keeps the Zn serum levels low. However, based upon studies in healthy subjects, neither the severe restriction of Zn in diet (e.g., <0.1-0.5 mg/day) is known to cause appreciable depletion in average serum Zn levels (upwards of 2 to 9 weeks are needed) nor the supplementation of Zn can restore the levels supposedly necessary to confer any benefit within the desired short window of 3-5 days of the crucial infection period (49). Usually upwards of 9-35 days are needed to bring the serum Zn levels back to basal levels even in individuals severely depleted of Zn levels (49). Furthermore, due to inter-individual variation in serum Zn levels and their response to Zn supplementation, serum Zn levels are never a reliable indicator of an individuals’ Zn status. The remarks made by Hess et al. 2007 based upon their analysis of nine intervention trials, summarizes the Zn serum levels status as “… an individual’s serum zinc concentration does not reliably predict that person’s response to zinc supplementation.” (whereas the) “Serum zinc concentration can be considered a useful biomarker of a population’s risk of zinc deficiency and response to zinc interventions, although it may not be a reliable indicator of individual zinc status.” (49, p-S403)

The variable(s) responsible for the observed association between Zn sufficiency and COVID-19 mortality remain undefined. We speculate that the increased mortality observed in high Zn sufficiency populations could be contributed by the existing underlying conditions of altered genetic composition, physiology, response to Zn supplementations/modulation in certain sections of the population [15-22,50]. It would be making them prone to display characteristic COVID-19 adverse illness as a result of increased oxygen radical generation or diminished oxidative stress handling system due to their physiologies’ propensity to promote the pro-oxidant role of Zn on supplementation (2). Furthermore, Zn overdoses would have the potential to negatively affect the activity of different Zn dependent antioxidative system components leading to oxidative stress and tissue damage, *e*.*g*., the R213G polymorphism of superoxide dismutase 3 (SOD3), a key member of the extracellular reactive oxygen species (ROS) neutralization system, found in 2-6 % of the population is known to make the R213G carrier population prone to oxidative stress promoted hypertension and increased risk of cardiovascular infractions and thrombosis (REFs in OMIM #185490 ‘SUPEROXIDE DISMUTASE 3; SOD3’ https://www.omim.org/entry/185490; additional info at http://www.wikigenes.org/e/gene/e/6649.html) - some conditions frequently associated with COVID-19 adverse events (51). Moreover, the Zn overdose is not uncommon in the highly impacted aged population of the developed countries that are more receptive to supplementation with or without a prescription (52).

Recently published meta-analyses of drugs being prescribed worldwide as a standard-of-care and the investigational drugs being evaluated in clinical trials for COVID-19 treatment, starkly indicate that the most promising drug for alleviating COVID-19 symptoms and decreasing the mortality had been such corticosteroids alone (53,54) that also suppress the Zn levels (55-57) along with increasing levels of oxidative damage protecting metallothioneins (58-59). A similar increase in metallothionein levels had been reported for 1,25-dihydroxy vitamin D3 (60) as well, another potential protective variable for COVID-19 (29). This serum Zn level suppression property of the select corticosteroids is also shared by steroid hormone estradiols which are naturally higher in females and youngs – two broad groups that had also consistently displayed lower vulnerability to COVID-19 adverse events (23). Whether these occurrences are coincidences or have mechanistic underpinnings only future investigations would reveal.

In conclusion, the observations presented, though circumstantial, underscore the need to revisit existing Zn supplementation guidelines for COVID-19 patients. Exploratory studies should be undertaken to correctly identify different underlying comorbid conditions that could make certain Zn supplemented individuals prone to COVID-19 mortality. The outcomes could help devise more comprehensive Zn supplementation practices for minimizing COVID-19 mortality among the vulnerable.

## Data Availability

All data are available in the manuscript/references provided.

## Ethical statement

The study is compliant with ethical standards. Considering the design of the study no human or animal rights were infringed upon.

## Informed consent

Considering the design of the study no informed consent was necessary.

## Figure Legends

**Supplementary Figure 1.**
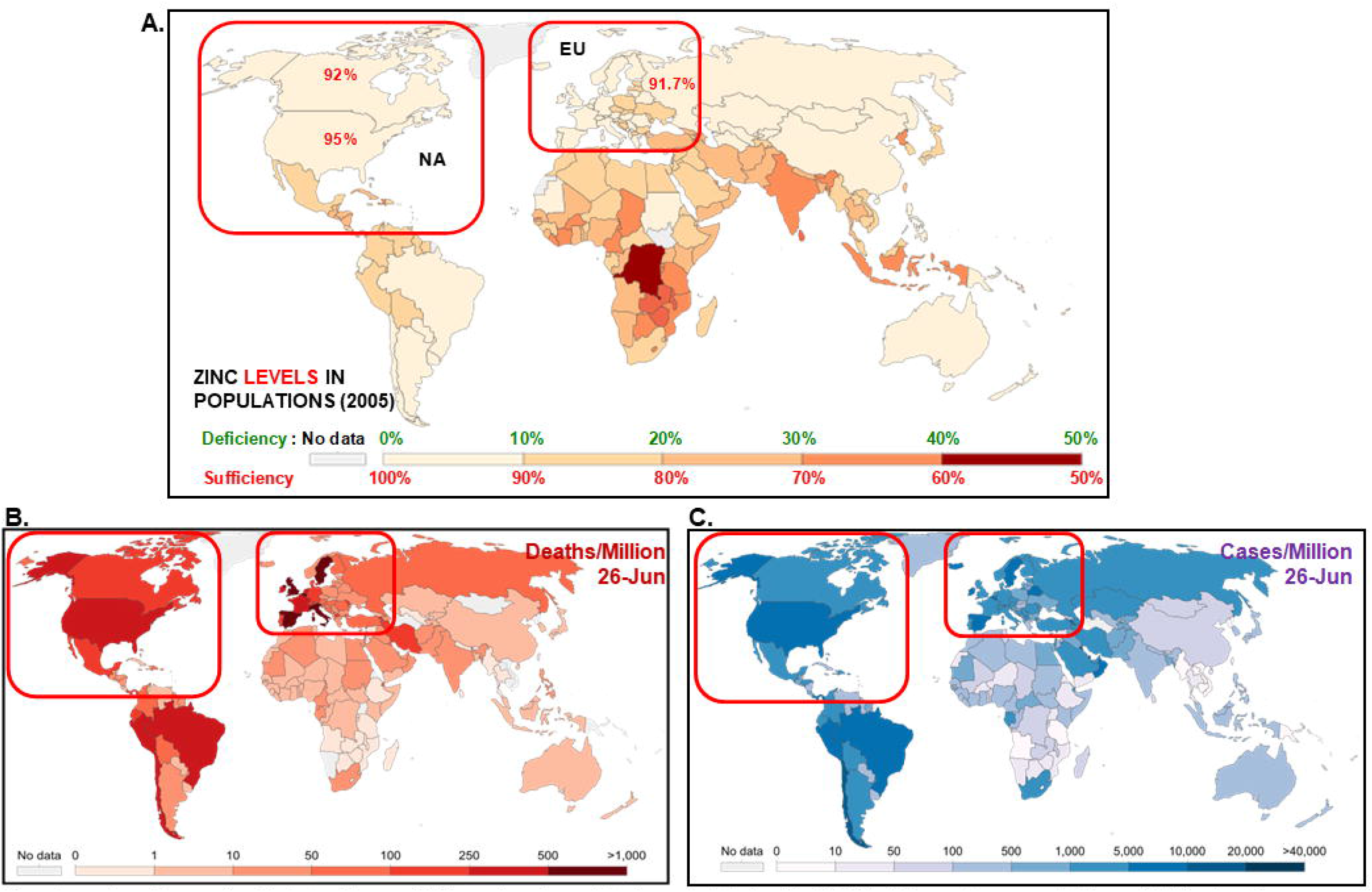
Globally, Zinc sufficiency levels are positively associated with COVID-19 impact on populations. **(A)** Zinc levels vary across countries. European (EU) and North American (NA) countries (red box) with higher development index, more diversified diet, with food fortification in place had higher Zn sufficiency. **(B)** The mortality per million population in socially similar EU and NA countries at a similar stage of 1^st^ wave strongly covary with Zn sufficiency (higher sufficiency-higher deaths). *For analysis of European nations see the main manuscript*. **(C)** Cases per million across countries. Images adapted from refs. 14, 25

**Supplementary Figure 2.**
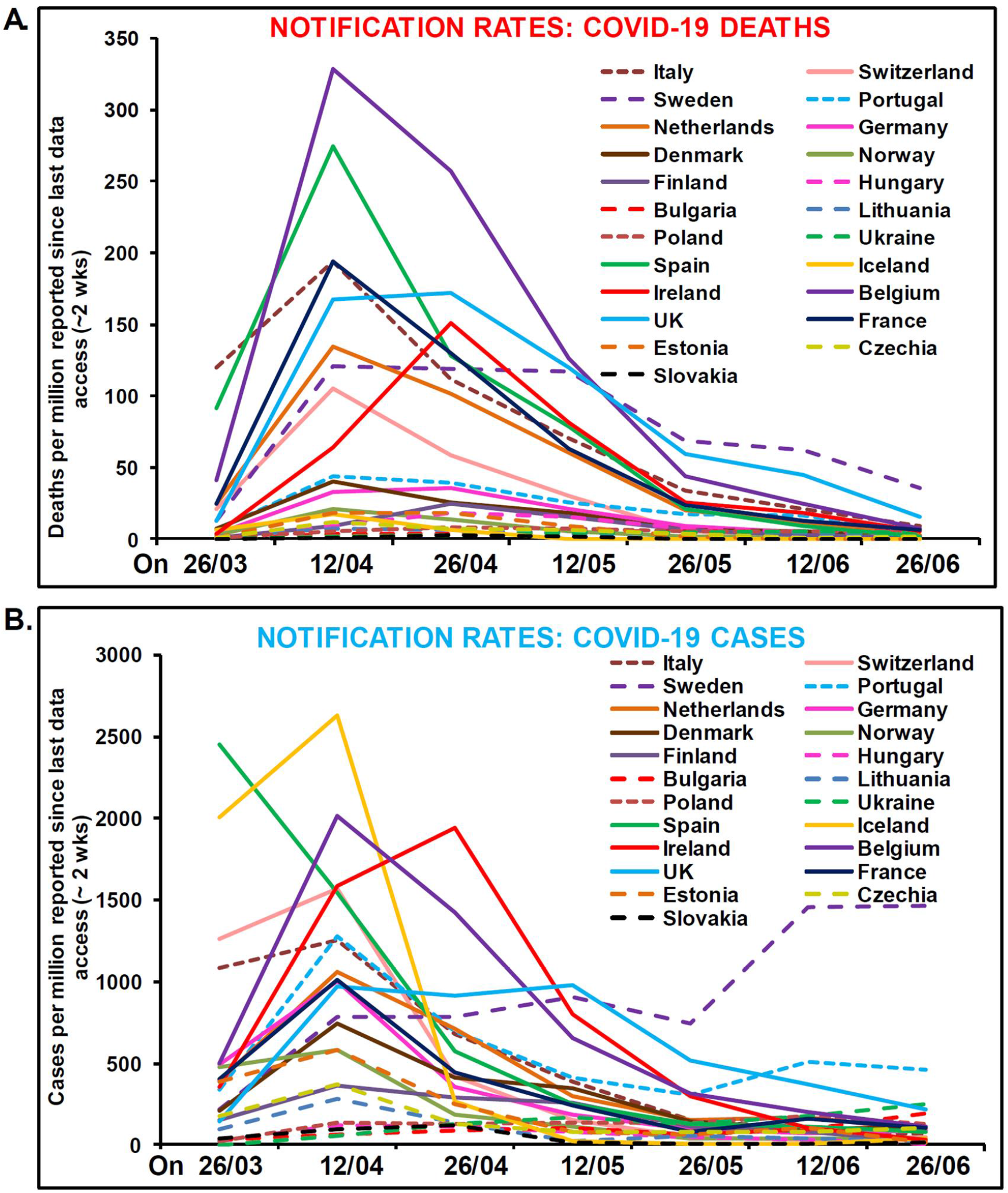
The wave of COVID-19 deaths (8 out of 10 were elderly) **(A)** and incidences (majority younger – without dysregulated Zn homeostasis) **(B)** as it traversed the countries analyzed in the current study

**Supplementary Table 1. COVID-19 incidences and mortality in socially similar European populations up to 26 August 2020**

